# Ecological impacts of climate change will transform public health priorities for zoonotic and vector-borne disease

**DOI:** 10.1101/2024.02.09.24302575

**Authors:** David W. Redding, Rory Gibb, Kate E. Jones

## Abstract

Climate change impacts on zoonotic/vector-borne diseases pose significant threats to humanity^1^ but these links are, in general, poorly understood^2^. Here, we project present and future geographical risk patterns for 141 infectious agents to understand likely climate change impacts, by integrating ecological models of infection hazard (climate-driven host/vector distributions and dispersal^3,4^) with exposure (human populations) and vulnerability (poverty prevalence). Projections until 2050, under a medium climate change (Representative Concentration Pathway (RCP) 4.5), show a 9.6% mean increase in endemic area size for zoonotic/vector-borne diseases globally (n=101), with expansions common across continents and priority pathogen groups. Range shifts of host and vector animal species appear to drive higher disease risk for many areas near the poles by 2050 and beyond. Projections using lower climate change scenarios (RCP 2.6 & 4.5) indicated similar or slightly worse future population exposure trends than higher scenarios (RCP 6.0 & 8.5), possibly due to host and vector species being unable to track faster climatic changes. Socioeconomic development trajectories, Shared Socioeconomic Pathways (SSPs), mediate future risk through a combination of climate and demographic change, which will disrupt current, regional patterns of disease burden. Overall, our study suggests that climate change will likely exacerbate global animal-borne disease risk, emphasising the need to consider climate change as a health threat.

**One Sentence Summary:** Climate change and socio-economic development dictate future geographical areas at risk of zoonotic and vector-borne diseases.

## Main Text

Climate change will profoundly affect the health and wellbeing of human societies^1^. Alongside direct impacts arising from physical processes such as sea level rise and extreme hydrometeorological events^2^, many indirect impacts to health will be mediated through changes in biodiversity, ecological communities, and associated ecosystem services^3^. In particular, climatic changes are reshaping the distribution and burden of zoonotic and vector-borne disease (ZVBD), as communities of hosts, vectors and pathogens are altering in abundance and geographical range in response to changing conditions^4,5^. Building resilience to these changes will require public health agencies to proactively build capacity to detect, respond to and treat ZVBDs as they emerge in novel settings, including in many regions with historically underdeveloped health systems^6^.

Biodiversity in general is expected to be negatively impacted by climate change (for many biomes and taxonomic groups^7–9)^, which contrasts with system-specific evidence that, of those examined, some key ZVBDs appear to be increasing in incidence or geographical area^10–12^. For instance, there is an emerging multi-model consensus that high burden diseases including malaria and dengue, will undergo significant increases and/or geographical redistribution in the coming decades, driven by climatic and socio-economic factors^13–15^. However, the net effects of climate change on disease risk and burden across the full breadth of vector-borne and zoonotic pathogens are not well understood.

Accurate prediction of animal-to-human infection (*‘spillover’*) patterns in space and time, and in response to climate change, ideally requires models fitted to fine-scale, systematic pathogen surveillance data from people and animals. However, most wildlife-associated ZVBD infections are neglected and data-deficient, which fundamentally limits such fine-scale prediction to all but a handful of diseases^16^. Statistical risk mapping approaches used to geographically interpolate present-day spillover risk (based on outbreak occurrence data) are unsuitable for longer-range projections due to the confounding effects of poorly-understood detection biases^17^. Models based on core ecological principles are a more appropriate basis for predicting future populations at risk of exposure to under-studied zoonotic pathogens^18^. The fundamental mechanism that constrains the local geographical hazard for wildlife-associated ZVBD infections, most of which have low human-to-human transmissibility, is the presence and abundance of competent hosts and vectors (i.e. a suitable host community^19^). Recent studies have shown that environmentally-driven predictive models incorporating climate information, based on host and vector distributions and movement, can accurately determine the broad geographical extent and limits of transmission for particular animal-borne pathogens^20–22^. Beneficially, such models can draw on large and well-established databases and models for biodiversity range dynamics^23^, host dispersal^24^ and known host-pathogen and host-vector associations^25^, to examine questions across a range of animal-borne diseases. As such, this approach provides a good framework to study future climate-driven trends in ZVBD endemic range dynamics, identify regions at high risk of emergence of multiple known pathogens, and understand how these intersect with changing socioeconomic vulnerability to capture changing public health priorities.

Here, we project effects of climate change impacts on the size and location of endemic areas of animal hosted or vectored infectious agents. We employ a Hazard-Exposure-Vulnerability (HEV) risk framework^26^ to generalise an ecological-epidemiological modelling approach of host and vector environmental suitability and dispersal (’hazard’) for 165 animal-borne or vectored infectious agents (114 viruses,18 bacteria, 30 endoparasites and 3 fungi) with present day and projected future human populations (‘exposure’), under 15 combined climate-socioeconomic (‘vulnerability’) scenario pathways from 2030 to 2080^27^ (fig.1, Methods). We collated all available data on associations between each agent and its currently-known hosts and vectors (i.e. representing the current state of knowledge, rather than a comprehensive understanding^28^) and then searched for information, for each agent, as to whether there was evidence of clinical disease in humans, recording countries that report infections, and more specific location where available (Data S1). We project the present (set at 2015) and future (i.e., for years 2030, 2050, 2070, 2080) geographic range of all hosts and vectors (n=1058 across all agents), using species distribution models, filtered by suitable land-cover using an area of habitat (AOH) approach^29^, and biologically realistic models of species dispersal^24^ to account for temporal colonisation dynamics (fig. 1). We then predicted the endemic geographical limits for each agent (henceforth *‘endemic area’*) by combining the predicted animal distribution data with a mathematical function determined by each agent’s dominant known transmission pathway to humans (88 were host only, 53 host and vector, 24 were vector only; across these groups 13 also included an intermediate or amplification step and 6 had domestic animals as a key transmission component). We validated the predictive accuracy of present-day endemic areas using the country-, or state-level infection reports (Data S1, S2), via sets of randomised pseudo-presence and pseudo-absence points within and outside the reporting area, using the area under the receiver operating curve (AUC) statistic. Once validated, we calculated the change in each agent’s endemic area across continents and among high priority infectious agent groups (e.g. WHO priority diseases, Data S1). To understand the drivers behind any trends, we then calculated the correlation between predicted endemic area size and future year for each agent, across all sets of future scenarios, and compared this to a randomised, null distribution of correlation values for the same sample sizes (Methods). We then examined the direction and size of endemic area change and mapped those areas most impacted by climate-driven changes in risk.

**Figure 1.**
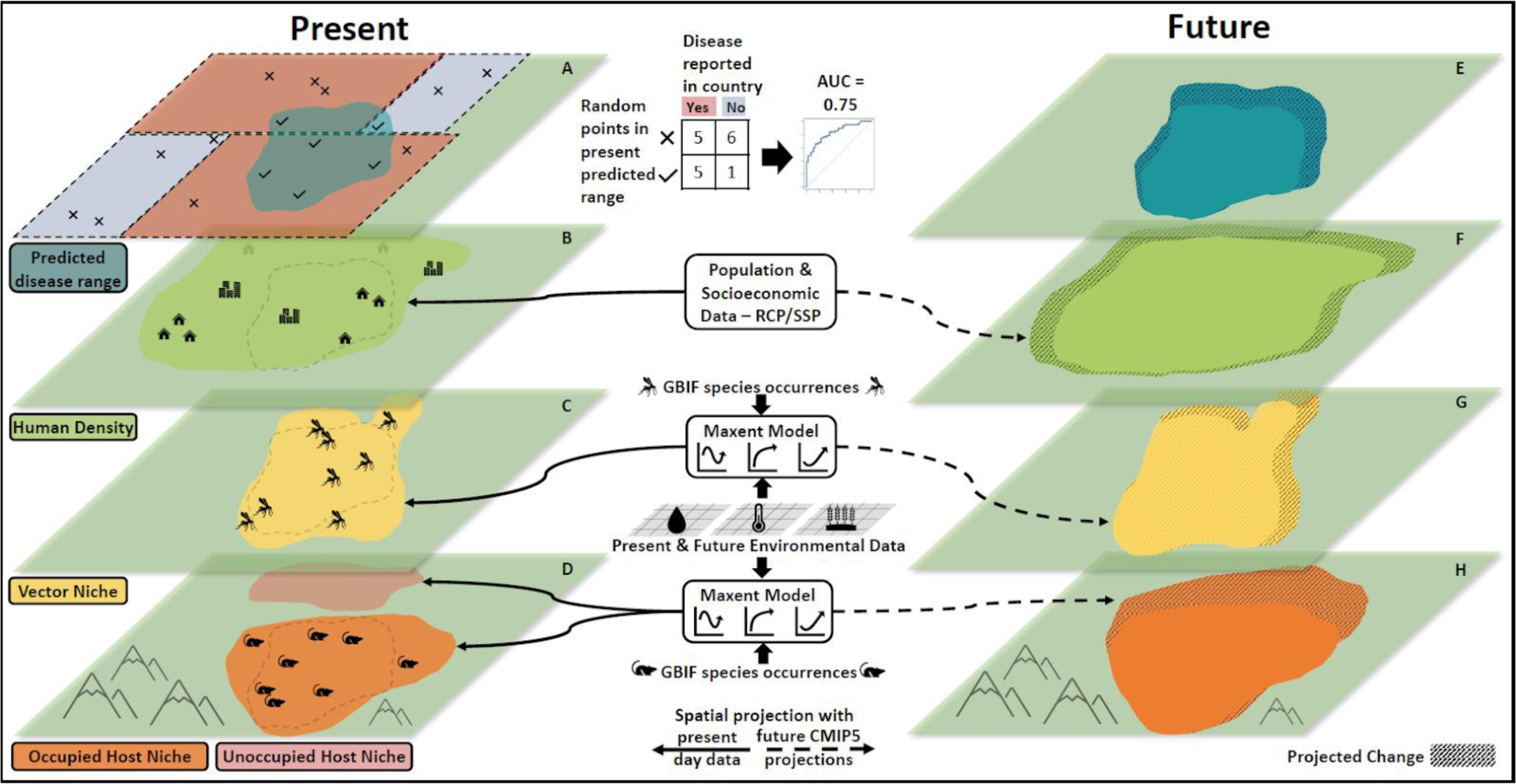
Diagram of present day and future projections of a disease endemic area. Panel A illustrates the predicted current disease endemic area (turquoise polygon) that is formed by the intersection of data within Panels B-D. Beige colour in A indicates countries with reported cases, while light blue shows non-reporting areas. Crosses and ticks represent points within and outside the endemic area, respectively, used to evaluate the model’s accuracy against country reports. Panel B displays human population density. Panel C & D represents a spatial occurrence model for vectors and hosts respectively, inferred using climate, and land-use information. Both panels exclude those areas without observed occurrences despite suitable climatic conditions (brown shape in D). Panels E-H project future scenarios: Panel E predicts future endemic areas by integrating data from Panels F-H, which use CMIP5 climate projections to forecast changes in vector (G) and host occurrence (H) and Shared Socioeconomic Pathways (SSP) to predict human populations (F). Dotted regions in Panels G and H signify potential future spread, constrained by biologically plausible movement rates derived from empirical studies.

Of the 165 animal-hosted or vectored infectious agents we examined, 141 (85.9%) had their ecologically modelled endemic area predictions matched to their present-day known endemic countries, or states, with an accuracy better than null expectation (i.e. AUC >0.5; 74.8% had AUC accuracy >0.6; 52.6% AUC>0.7). For those agents for which fine-scale validation data were also available (Data S3), our validation returned a mean accuracy again above random expectation (mean AUC=0.682, sd=0.172, n=10). For only 101 of these agents, however, we could confirm clinical evidence of disease in humans (all results hereon reported using this group, termed ZVBDs), and for this group by 2050, there was an overall 9.6% (bootstrapped 95% confidence intervals (CI) 6.9-15.8%) increase in mean area at risk. This pattern was broadly similar across continents (fig. 2a). The ZVBDs within global priority groups all showed mean expansions (fig. 2b), with National Institute of Allergy and Infectious Diseases (NIAID) group C being the only group that had CIs that crossed zero. Surprisingly, endemic area growth trends to 2080 became weaker with increasing levels of warming (i.e. highest risk increases under the lowest radiative forcing pathway of RCP2.6; fig. 2c), supporting recent work projecting changes in potential rates of cross-species viral transmission across different emission scenarios^5^. This may be because, in higher-warming scenarios, animal species are not able to disperse quickly enough to track the rapid movement of their preferred environmental niche^5,24^, a finding again consistent with expectations of greater biodiversity loss under more severe climate change^30^.

**Figure 2.**
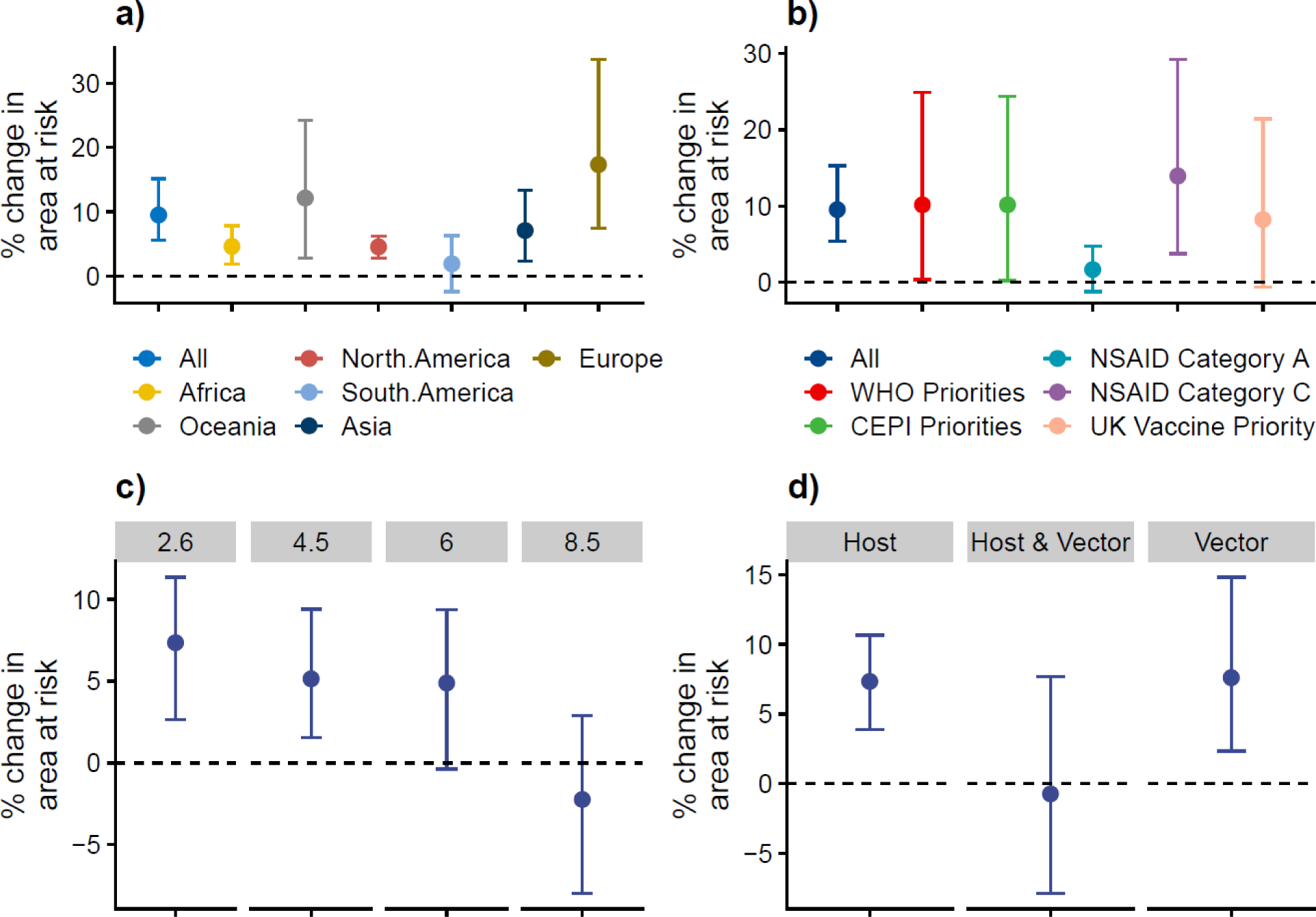
Mean change in geographical area at risk of wildlife- and vector-borne disease under climate change. Dots are mean values and whiskers the 95% CI across diseases in the sample. Panels show mean and standard error of the per disease area change a) in all continents, and within individual continents for the year 2050; b) for all diseases and for groups of priority diseases for the year 2050, c) across four RCP pathways (2.6, 4.5, 6.0, 8.5) for the year 2080, d) for groups of diseases with different transmission mechanisms for the year 2080.

Lastly, ZVBDs with more complex transmission pathways (involving both wildlife host and vector species) had, by 2080, a range reduction compared to diseases that were transmitted just by vectors or from wildlife. This suggests that more complex infection systems (i.e. transmission requiring co-occurrence of at least two quite unrelated species) may be more susceptible to ecological disruption by climate change^5^. Indeed, organisms show contrasting responses to climate change^31^ and disease systems that require several co-occurring species within the transmission cycle and for spillover to occur^32^ may see species moving in opposing directions or at different rates, reducing the geographical range within which transmission can occur, in the absence of adaptive responses.

To understand how consistent ZVBDs were in their climate change response, we then examined the frequency of expansions compared to reductions. By 2080, more than half of ZVBDs saw an expansion (54.5%) of their endemic area over time with climate change, while 27.4% remained similar-sized and 18.1% contracted, a finding that contrasts the widely accepted negative impacts of climate change on biodiversity more generally^7,33^, and similar region-or taxon-specific work (e.g. more than 50% of birds in Australia show projected endemic range contractions with climate change^34^). This pattern of more expansions than expected held when drawing different sized bootstrap samples of the 101 ZVBDs (fig. S1). A higher proportion of endemic area expansions (fig. S2) were projected for vector-only agents (e.g. dengue; median change in endemic area of 28,343km^2^ per year), compared to agents with non-vector-borne transmission (i.e. host to humans through direct or environmentally-mediated routes). This again contrasts with agents whose sylvatic and spillover cycles involve both host and vector species (e.g. Lyme disease, plague), which show more contractions than expected under the null (fig. S2) and a median endemic area decline of −2664 km^2^ per year. The large expansions for vector-borne pathogens are consistent with previous research that describes a positive association of climate change and disease risk^10,11,35^, often attributed to physiological drivers such as a tight coupling between ambient temperature and vector biological and life history parameters^12^.

Next, we examined trends in the directionality of endemic area shifts, which could inform priorities for emerging disease surveillance, by examining the expected change in location of endemic area centroids by 2050 (fig. 3a). We project consistent poleward range shifts of endemic areas in temperate regions, across ZVBDs (1.182 times more northward shifts in the northern hemisphere than expected, and 2.54 times more southward shifts in the southern hemisphere, χ-squared = 17.405, df = 3, p-value < 0.001) (fig. S3). ZVBDs in northern temperate areas (>23.5°N) have a mean centroid shift of 0.87°N (96.64km at the equator; 81.97-111.38km 95%CI) from 2030 to 2080, whereas those diseases in southern temperate climates (< −23.5°) have a mean shift of 0.65°S (72.264km at the equator; 31.65-112.87km 95%CI) in the same period. Tropical areas are consistent with temperate areas in terms of the direction of mean shift, but show less consistent patterns, with smaller overall distances (0.16°N and 0.48°S for north and south tropics respectively). Indeed, there is a wide variety of projected endemic area shift responses in the tropics, with northern tropical areas having 1.61 times and southern tropical areas 1.28 times more equatorial movements than expected (χ-squared = 17.405, df = 3, p-value < 0.001). Overall, there appears a general pattern of increasingly larger range shifts towards the poles (fig. S4), meaning that more northern and southern areas will need to adapt quicker to patterns of changing disease risk.

**Figure 3.**
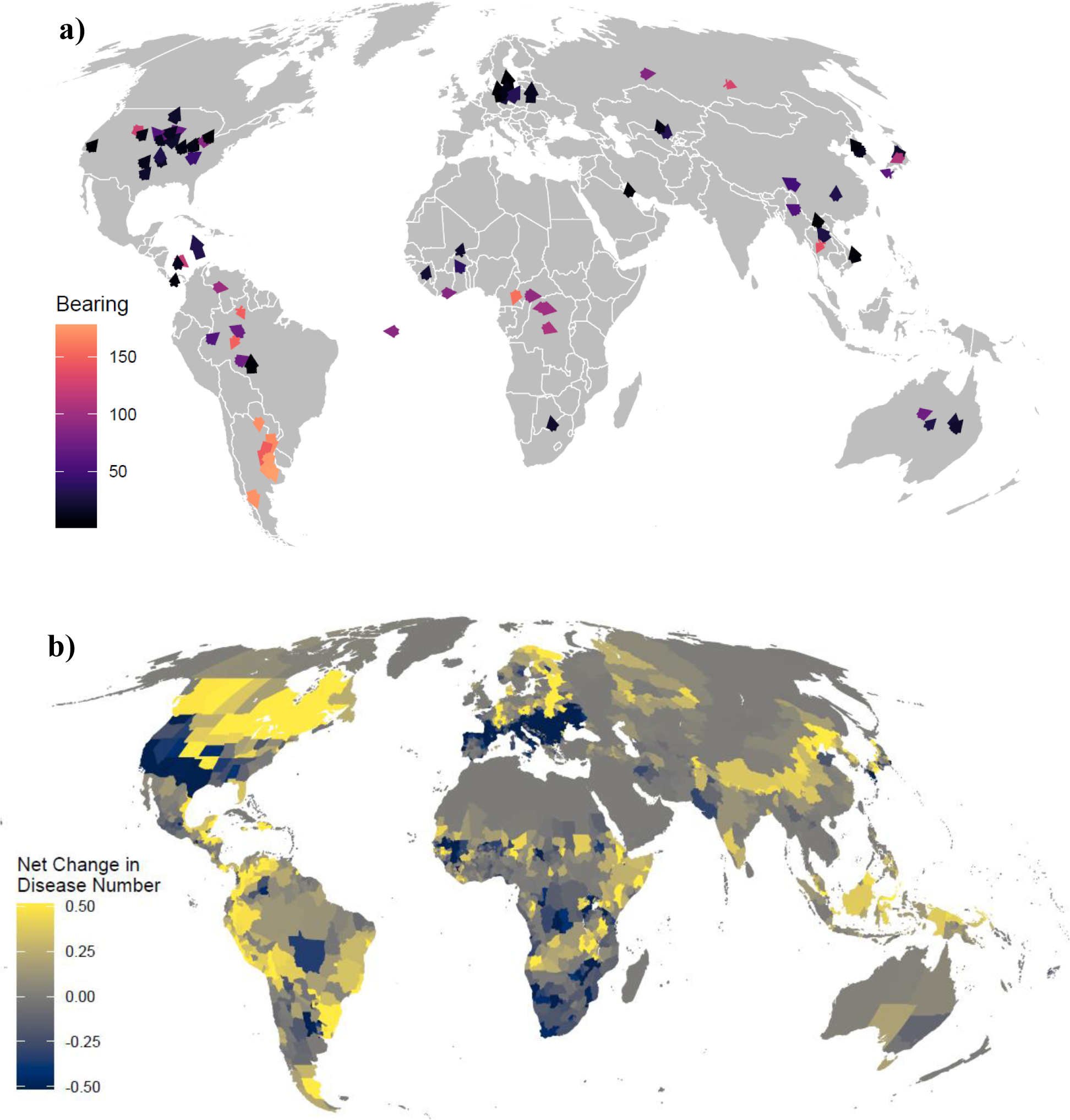
Global maps of disease endemic area range shifts. Panel a) represents present location of disease endemic area centroid, marked by arrows, colour coded by absolute bearing to reinforce the direction of the arrows, which point in the direction of future shifts caused by climate change by 2050, for 101 animal and vector-borne diseases; and b) how the movements of these risk areas alters the net mean per pixel disease richness within administrative level three^40^ regions for these same set of diseases by 2050. Projection is Mollweide and units are degrees of latitude and longitude.

To understand how these geographical shifts may translate to net impacts on disease burden, we mapped the net changes to ZVBD endemic areas to 2050 under a medium-change scenario (RCP 4.5 SSP2 – fig. 3b). There is evidence of geographical foci with moderate-to-high net increases in ZVBDs over the next 30 years relative to the present day, such as north-eastern North America, northern Scandinavia, the Horn of Africa and the southern edge of the Tibetan Plateau (fig. 4). In Southern Europe, southern United States and central Africa, however, there appears some consensus for a decrease in the number of ZVBDs, possibly due to climate change reducing environmental suitability for the current host species that live there, or due to knowledge gaps about alternative (as-yet-unknown) ZVBDs that might replace them. Elsewhere, principally the tropics, there are areas of increasing and decreasing richness in close proximity, suggesting there is little clear evidence for overall geographical patterns of change in ZVBD diversity, at least at this coarse spatial scale (Data S4). For much of the world, however, the relatively small set of ZVBDs included in this analysis (101 compared to the estimated 1407 zoonoses^36^) means there is incomplete information to uncover any trend (i.e. areas shown in light grey in fig. 3b).

**Figure 4.**
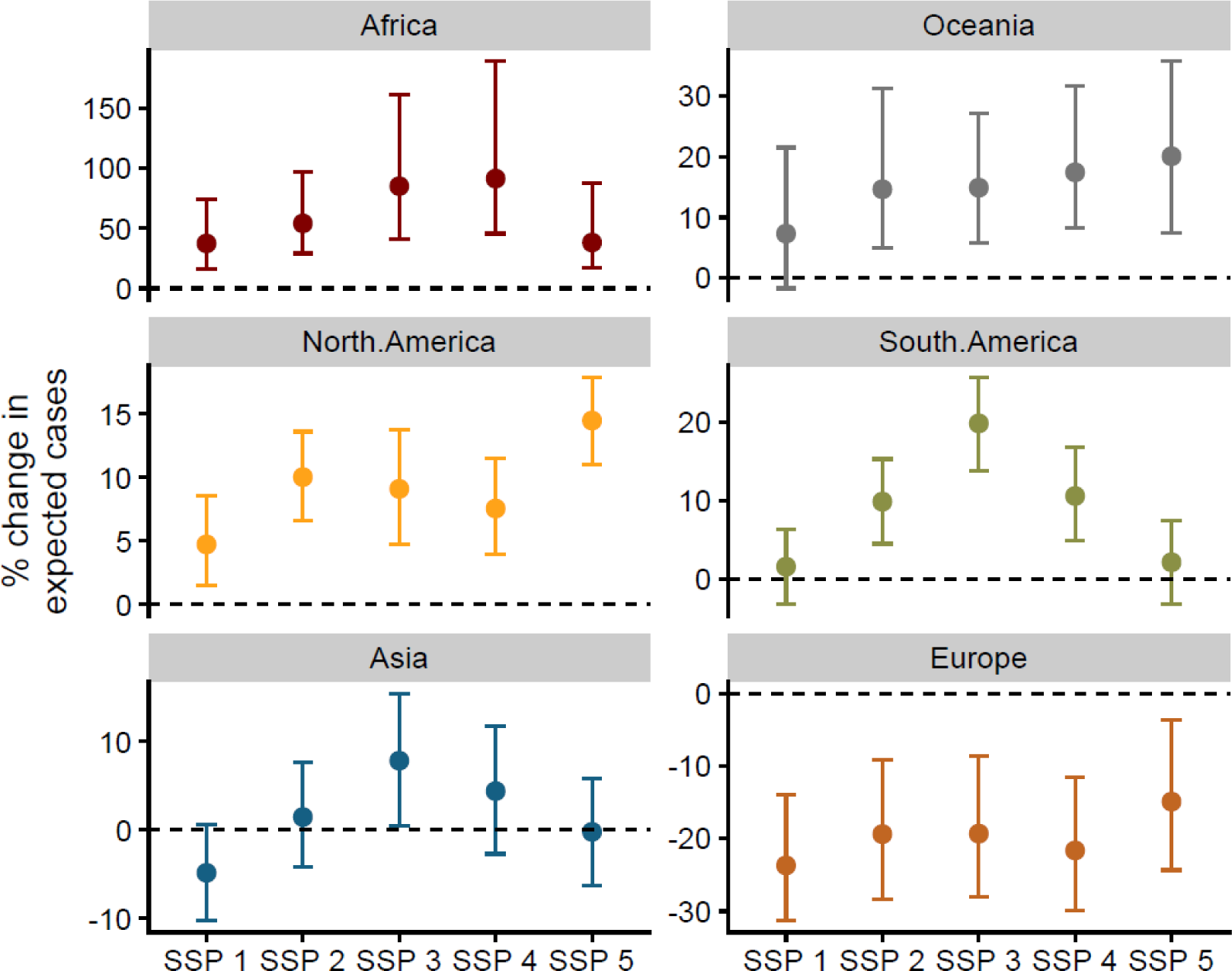
Percentage change in expected number of wildlife- and vector-borne disease exposures in economically vulnerable populations (those earning less than the 20th percentile of income) continentally, by 2050, across 5 different shared socio-economic pathways (SSP). Dots are mean and whiskers 95% CI. Where SSP1 - the focus is on sustainable development; SSP2 - where trends from the past continue without significant efforts to address sustainability or inequality; SSP3 - a future marked by high regional competition, economic inequality, and slow technological progress; SSP4 - explores a future characterised by high levels of socioeconomic inequality; SSP 5 - emphasis is on high energy demand, rapid economic growth, and a reliance on fossil fuels.

Lastly, to estimate how these geographical shifts in ecological suitability may translate to a net change in human health impacts, we projected risk across five socioeconomic development scenarios (SSPs – outlined in fig. 4 legend), using a HEV framework i.e. Risk = Hazard * Exposure * Vulnerability. Risk, or expected exposures, per grid cell, per ZVBD was the product of *Hazard*, defined as a value of 1 if a cell is located within the modelled endemic area and 0 if not; *Exposure,* defined as the predicted human population in the grid cell^27^; and *Vulnerability,* the fraction of people in relative poverty (set as the proportion of people earning less than the 25th percentile of local income range). Our projections assumed that human contact rates with hosts and vectors are a linear function of human density, and that the proportion of low earners is an effective index of the factors that increase vulnerability to infection hazards, such as lower health systems access and poorer nutrition, housing and sanitation infrastructure. This assumption is generally empirically supported; poverty correlates with worse disease outcomes, higher incidence and greater impacts on future livelihoods^37^.

Our models project strong regional differences in expected burden change (fig. 4), with most continents, except Europe, seeing consistent increases in burden due to the combined effects of climate change and population growth. Africa had by far the largest increases in expected burden (between 25 and 75%) compared to smaller increases in Oceania, North America and South America (between 5 and 20%). South America and Asia see larger increases for SSP3 (a ‘regional rivalry’ scenario with poor international cooperation on climate mitigation, adaptation and development), compared to more optimistic SSPs, such as the climate-focused cooperation proposed in SSP1, or technological-solution focus in SSP5^27^. Europe, in contrast to other areas, is projected to see a reduction in expected burden across all SSPs, likely due to the projected reductions in ZVBD richness being concentrated in the currently most populated areas, and the increasing richness in the less densely populated regions, though the true pattern is potentially confounded by limited knowledge, or ineffective modelling, of invading diseases. In summary, ZVBDs in Africa are less sensitive to SSP scenarios (fig. S5, S6) and more strongly impacted by climate change, indicating that the general pattern of endemic area expansion is causing novel disease to spread into already highly populated areas, while those in Asia and Europe are more sensitive to patterns of population growth or reduction, as climate change drives spread generally into currently less populated areas e.g. boreal forests of Scandinavia and the Tibetan plateau (fig. 3).

Taken together, the climate change effects on the ecological niche processes underlying the distribution of animal host species appear to be both overall increasing the size and shifting the distribution of risk areas for wildlife-borne diseases. The modelling approach we use means that these projections are broad-scale indicators of underlying trends; due to data limitations our models cannot represent the numerous smaller-scale social and ecological processes that drive fine spatial and temporal patterns of transmission infection, nor any counteracting effects of pathogen and host/vector adaptation to changing conditions. Our results however are indicative of the large-scale emerging patterns of climate change pressures on wildlife-borne disease, and these general trends suggest pathogen movement into areas with potentially immunologically-naive human populations, possibly altering large outbreak and pandemic risk^38^. Furthermore, this means the current type, capacity and location of healthcare, diagnostic and urban infrastructure may not be appropriate to mitigate the changing patterns of risk seen in the near future (e.g. open gutters on housing given shifting dengue risk^39^). Collectively, humanity needs to respond to the disruption of climate change to prepare societies for future increases in zoonotic and vector-borne disease burden.

## Data Availability

All data produced in the present study are available upon reasonable request to the authors

https://www.dropbox.com/scl/fo/lrsts3ipcalppzktlol9e/h?rlkey=dnvh4rgpbe225pxdwwekpwq4z&dl=0

## Acknowledgments

We thank Roi Maor for help with the database.

## Funding

This research was supported by an MRC UKRI/Rutherford Fellowship (MR/R02491X/1, MR/R02491X/2), Sir Henry Dale Research Fellowship (funded by the Wellcome Trust and the Royal Society) (220179/Z/20/Z, 220179/A/20/Z) (DWR), and The Trinity Challenge Sentinel Forecasting Project (KEJ, RG).

## Author contributions

Conceptualization: DR, KE Methodology: DR, RG Investigation: DR Visualisation: DR, KE, RG Funding acquisition: DR, KE Project administration: DR Writing: DR, RG, KE

## Competing interests

Authors declare that they have no competing interests.

## Data and materials availability

All input data, code and summaries from the model runs to run models are available at the following URL: https://www.dropbox.com/scl/fo/lrsts3ipcalppzktlol9e/h?rlkey=dnvh4rgpbe225pxdwwekpwq4z&dl=0

## Materials and Methods

Our modelling approach consisted of five main steps, as summarised in fig. 1: First, we collated data on all infectious agent-by-host species interactions where wild animals were involved in the transmission pathway, noting that this work pre-dates and complements other recently published datasets^41^. Second, we collated occurrence data for all non-human actors in the transmission process, created individual species distribution models (SDMs) for each host and vector species, and then used these models to predict gridded layers of geographic suitability using both present and future climatic conditions. Third, we created an infectious agent spatial occurrence suitability composite layer for present day, by combining suitability for all transmission components mathematically, evaluated the predictive performance of this resulting layer against spatial data on infection, and then used optimised presence-absence thresholds from the validation process to threshold suitability and delineate the present-day infectious agent endemic area predictions. Fourth, we generated the same composite layers using future host and vector projections under climate change, and used the per-agent threshold values to determine whether an agent’s endemic area was projected to expand or reduce. Fifth, we used a contact rate model to examine how changes to agent’s endemic areas interact with projected changing human populations under a variety of future socio-economic development scenarios to establish a compound risk value. We expand on these sections below.

### Host-vector-agent data collection

Data on infectious agents, host and vector associations were collated from the literature (Extended data table 1) for zoonotic or vector borne infectious agents found in wild animals and have potential for infecting humans (data collation was undertaken over years 2014-2020). To structure the search, we started with all the named infectious agents with a wild animal (host/vector) involved in the transmission pathway, that were listed in a review of human infectious diseases for which there is spatial occurrence information^42^ available. This set was chosen as a) it contains the vast majority of high-burden animal-borne disease, and b) the spatial data in this publication can provide a basis for validation of the modelling outputs. For all the zoonoses in this dataset we searched the literature to identify the putative reservoir host, i.e. species that carry an infectious agent with minimal cost to individual fitness and that play a key role in maintaining infection across the landscape. For agents transmitted by vectors, where possible, we identified principal vectors rather than rare or occasional ones. We focused on putative reservoirs vectors rather than incidental actors, where indicative data were available, as these species are most likely to have an impact on the distributions of infection risk, if they move in response to environmental change. For many infectious agents, there has been little review or critical analysis of principal hosts or vectors, and for a few agents we identified host/vectors for only one part (e.g. continent) of the endemic geographical range. Indeed, since the highest impact diseases are generally much better-studied, our attempt to create a comprehensive dataset will suffer from many systematic biases. For instance, more information will be weighted towards agents that directly affect populations of richer industrialised countries as they tend to spend more funding on research. Furthermore, certain groups of agents that have caused large, disruptive outbreaks will also be more researched and subsequently better understood^28,43^. Our analyses attempted to adjust for these biases through weighted bootstrapping of the data when visualising and summarising.

To validate geographical predictive performance of models, we recorded the countries which have reported each agent (Data S1). For infectious agents documented in two countries or fewer, where country boundaries could grossly overrepresent the size of the endemic area, we undertook a literature search for locations where cases had occurred. We used those locations to assign as “present”, all administrative level 3 geopolitical entities^40^ in which cases for that agent occurred (Data S2). For a subset of agents (n=10) there were geolocated cases or outbreaks available from online databases to provide a more detailed, fine-scale validation of model outputs (Data S3).

### Mapping ecological hazard using hosts and vector distribution

We used an Area of Habitat (AOH) approach^29^ to ensure that locations where host and vector species were deemed present, or dispersed into according to climate based models, contained habitat suitable for those species. We first determined habitat suitability for each species, based on land-cover-land-use, across 7 broad categories (Primary – forest, Primary – non-forest, Secondary – forest, Secondary – non-forest, Pasture, Cropland and Urban)^44^. For vertebrates, these categories were then matched to broad IUCN habitat preference data at species-level, which provides three levels of suitability for each species; we gave a value of 1 to suitable habitat, a value of 0.5 to marginal habitat and a value of 0 to unsuitable habitats. We multiplied the resulting climate suitability map by the climatic niche prediction for each species (as defined below) to produce an area of habitat species occurrence layer^29^. For invertebrates, where there is limited information on habitat type, we used occurrence point data to determine whether habitats were suitable or not. We did not alter the habitat layer in the AOH calculations for future predictions because there is extremely wide uncertainty in future modelled trajectories of land change^45^, which are highly sensitive to difficult-to-forecast factors such as sociopolitical change, patterns of market demand and unexpected global shocks, as well as model assumptions.

Although a limited selection of future land use scenarios are available for global change modelling (e.g. Land Use Harmonization), unfortunately, these do not provide a means to quantify uncertainty in future risk associated with future land change patterns. We therefore focused on climate change as the key driver in this study, where the availability of multiple general circulation models (GCMs) enables a robust quantification of uncertainty in forecasts.

Next we constructed potential climatic niches for all host and vector species identified above using the MAXENT^46^ algorithm, with rasters representing all 19 BIOCLIM climatology variables and altitude^47^ as input variables. We used the MAXENT algorithm because it performs accurately for many datasets and is both computationally efficient and has a very high capacity for automation^48^. For presence points in the MAXENT inference, we downloaded occurrence data for hosts and vectors of each of the 586 identified agent-animal associations, using the API of the online database Global Biodiversity Information Facility (GBIF; using the *spocc* function spocc package^49^). Specifically, for each taxon we checked for misspelling or out-dated names against a set of existing taxonomies using the taxize package^50^ and then downloaded all recognised synonyms (*synonyms* function taxize package^50^). We then downloaded all geolocated occurrence data for all taxa and all their species level synonyms. For each infectious agent we choose subsets of those occurrence points that were located in the global subregions^40^ that contained countries that had reported the agent^42^, such that even if occurrences of an agent-carrying species were located in Africa and Asia but autochthonous infection only known to take place from species’ individuals in Asia, we only used Asian occurrences, since sub-species can have difference climatic tolerances. For example, for Lassa fever virus, the host *Mastomys natalensis* occurs in a variety of habitats across the whole of sub-Saharan Africa but Lassa Fever cases are restricted solely to West Africa^21^ and, therefore, only *M. natalensis* occurrence points from West Africa were used for the host niche estimation for this agent. This means that widely occurring species hosting more than one agent could either have: More than one regional climate niche model inferred for each different agent they host, or a single model if the multiple agents they host all occur in the same subregion. However, if there were too few occurrence points within the endemic subregion (n<25) we then downloaded all species occurrence points in the encompassing region. If again there were too few points (n<25) we did not model this animal-agent interaction. This resulted in 390 animal-agent links being brought forward to the next stage of modelling.

Before inference, we verified and cleaned the occurrence data from GBIF. First, from all datasets we removed all points that were not on land or an island according to a high-resolution coastal boundary dataset (according to gsshs data set *wrld_simpl* function in *maptools*) or had errors in the associated data file. Second, we removed the most imprecise points i.e. with less than 2 decimal place precision. Third, to reduce the impact of any spatial bias of sampling points we removed any points that co-occurred in 1-degree grid cells globally. Then to infer the niche model we ran 50 iterations of the MAXENT algorithm using random subsets of each species’ occurrence dataset, with random regression model configurations and random numbers of pseudo-absence points from 100 to 5000 per sub-model. For input data we downloaded all 19 bioclimatic variables for present day (averages 1960-2010) and altitude in 30 second grid format^47^. Using 10% holdout samples to calculate AUC, and employing an ensemble prediction approach, as it is shown to be more effective than choosing a single best-performing model^51^, we combined the rasters of global spatial predictions of the top five best performing models creating a mean prediction value for each grid cell. We undertook this inference approach for all hosts and vectors for which there was available data, and then predicted for each species by agent combination a layer of mean climatic niche suitability (*predict* function from *raster*) for the present day.

For future niche predictions we used the same input data variables as for the Maxent inference (bioclim and altitude) which were downloaded for the individual years 2030, 2050, 2070 and 2080, based on general circulation model (GCM) future modelled climates from the Coupled Model Intercomparison Project 5 (CMIP5) (http://ccafs-climate.org/data_spatial_downscaling/).

These layers consisted of summaries of spatial predictions from 26 different climate models with variations (Data S5) under three different RCP scenarios (RCP 2.6, 4.6, 6.0) giving a total of 318 possible future scenarios over all four future years. Due to high computational costs, we sampled 14,625 species-by-model scenarios by choosing approximately 50 model by RCP scenarios at random for each of the 390 animal agent associations, representing 11.79% of the 124,020 possible combinations. This random subsampling of GCMs by species and year combinations was a compromise for computational time (i.e. predicting for all combinations of model, species and year would have been unfeasible) while ensuring that projections captured the full breadth of modelled climate futures across the ensemble of GCMs from CMIP5.

### Present day agent hazard, validation, thresholding

The step above produced predicted present and future raster layers for each agent by wild species interaction. Where present, multiple host and vector occurrences were combined, for each grid cell *i*, agent *j*, and time *t* (initially this was just for present day set as 2015), as follows:

Host Occurrence Probability (*P_H,i,j,t_*):

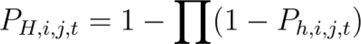

where *P_H,i,j,t_* is the probability of at least one host occurring in cell *i* for agent *j* at time *t*, and *P_H,i,j,t_* is the probability of occurrence for a putative host species in cell *i* for agent *j* at time *t*.

Vector Occurrence Probability (P*_v,i,j,t_*):

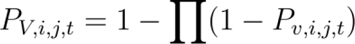

where P*_v,i,j,t_* is the probability of at least one vector occurring in cell *i* for agent *j* at time *t*, and *P_v,i,j,t_* is the probability of occurrence for a putative vector species in cell *i* for agent *j* at time *t*.

We then combined these species distribution grids into a single agent hazard layer using a mathematical function determined by the agent’s presumptive transmission pathway to predict the endemic geographical limits for each agent (henceforth *“endemic area”*):

Hazard Estimation (*H_i,j,t_*) for Host-to-Human Transmission:

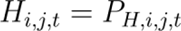

Hazard Estimation (*H_i,j,t_*) for Human-to-Vector-to-Human Transmission:

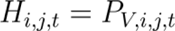

Hazard Estimation (*H_i,j,t_*) for Host-to-Vector-to-Human Transmission:

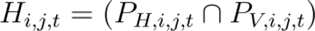

Hazard Estimation (*H_i,j,t_*) for Host-to-Human Transmission with Amplification Component:

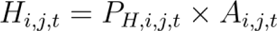

where (*A_i,j,t_*)is the density of amplifiers in cell *i* for agent *j* at time *t*.

Hazard Estimation (*H_i,j,t_*) for Host-to-Vector-to-Human Transmission with Amplification Component:

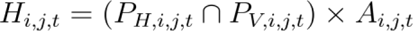

Applying the above equations resulted in a single present day hazard grid for each agent. We then validated these resulting hazard grids using data on known agent occurrence. Fine-scale data (i.e. point occurrences) on the known endemic area are available for a small proportion of agents (6%), so we needed to take a multi-tier validation process; firstly, at country level, secondly at state/county level for agents found in 2 countries or less, and lastly for the few diseases with geolocated case occurrence data available, we used point locations. We employed an area under the operating curve (AUC) algorithm to calculate predictive accuracy across these scales.

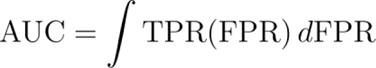

where TPR(FPR) represents the true positive rate (TPR) value corresponding to a given false positive rate (FPR) value. True Positive Rate (TPR) is defined as the ratio of true positives to the total number of positive points. False Positive Rate (FPR) is defined as the ratio of false positives to the total number of pseudo-absences.

For most diseases, where we had only country level infection data, during each run we generated random absence points (using 100, 500, 1500, 5000 or 10,000 points selected randomly) across the whole endemic sub-region and random presence points (i.e. 100, 500, 1500, 5000) within the boundaries of reporting countries. After validating, we then thresholded each hazard layer by optimising by AUC score. From 25 replicates of this validation, we created three threshold values: the minimum optimum value, the mean optimum value and maximum optimum value to act as a sensitivity test on this process. These optima gave qualitatively similar results (fig. S7; Data S6) and so we used the mean optimum hereon. Then taking all hazard layers, any grid cells with values below each of the mean threshold level were designated as not present (i.e. marked as NA) and then “presence” polygons were drawn around all contiguous positive grid cells i.e. designating those areas above the presence threshold. Then referring to the points downloaded from GBIF to create the presence models we only kept those “presence” polygons in the analysis that had known occurrences, to remove any suitable areas that had low evidence of ever being occupied.

### Future change in hazard

The future projected species occurrence rasters were then combined into single future hazard layers using the same equations as in above step, for each climate emissions scenario by year by climate model combination. One difference from the present day calculations described in the previous step, was that there was also variation by climate models, so we averaged across different future climate models for each climate emissions scenario (RCP 2.6, 4.5, 6.0) by decadal climatology (referred to as by the initial years 2030,2050, 2070,2080) to create 12 future hazard layers per agent. Next, using the threshold for each agent set in the previous step, we again designated any grid cells below the respective suitability threshold as ‘not present’, and those above the threshold ‘present’, to result in binary layers containing predicted future presence and absence.

To model biologically realistic species dispersal from the present-day range position to newly-suitable areas (i.e. range dynamics), we used the present day polygon as a range boundary to determine those ‘present’ cells that were occupied both presently and in the future. Then, for all cells outside the present day range, those suitable cells in the future predictions were only included as occupied in the future if they matched a set of diffusion criteria: Cells were only marked as present if they were less than 2 cells per decade from the present day range edge depending on the year (to match the empirically observed average dispersal rate from a large scale meta-analysis^24^), which meant change could only occur contiguously from the current range edge (so only known occupied areas were included in future predictions) and at a rate calculated using the empirical data estimate. All other cells, if there were any outside this area, were marked as not present. Therefore by 2080 each part of the range edge can only grow or be reduced by a maximum of 14 cells from its present-day location. Furthermore, we used present day land-cover type and habitat preferences of each species to prevent movement of animals into unsuitable habitat^53^ e.g. grassland species into forest. Finally, we then drew a polygon around all remaining contiguous grid cells and calculated future range size and centroid location for all 12 future layers for each agent.

We then, for each known infectious agent (n=101), calculated the correlation between (Spearman rank; function *cor*; base package in R^54^) the number of cells in the endemic area and year, using the years 2030-2080. We then repeated this, sampling different size subsets of the 101 agents (groups of 40, 56, 65, 71 sampled randomly 100 times) attempting to control for biases resulting from some geographical areas and some agent types being sampled more often, by weighting less represented agents by the reciprocal of the relative sample proportion (function *sample*; base package in R). For each sample we then randomised cell number (area) with respect to the year and took another correlation measure as a null expectation, to capture the frequency of strong, weak and neutral relationships that would be expected to be generated by chance using data of the same size and mean values. We then categorised all these correlations as either being negative (Rho < −0.25), neutral (Rho> −0.25 but Rho<0.25) or positive (Rho>0.25) and counted the number of samples where the cell number-year relationship fitted into these categories. We also repeated this process with a different threshold (i.e. Rho thresholds −0.5 and 0.5; −0.1 and 0.1) to test the sensitivity of the result to this value.

We then examined how the mean location of the endemic areas changed by measuring where their centroid moved from the year 2030 to 2080. We then labelled the agents depending on where the centroid was located latitudinally. The endemic areas in northern regions (>23.5°N) and those areas in southern regions (< −23.5°S) were designated as temperate, while all those in between were designated as tropical. For tropical endemic hazards we then designated between those that sat north of the equator (>0°) and those south (<0°).

### HEV Risk Model

Finally, we employed a Hazard-Exposure-Vulnerability (HEV) risk framework^26^ across agents, generalising an ecological-epidemiological modelling approach previously used to predict future Ebola and Lassa fever outbreak potential^20,21^, assuming homogeneous random contact within each grid cell. For each grid cell *i,* per agent *j,* at each year time point *t*:

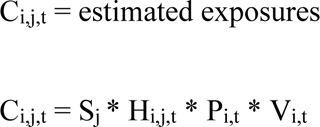

where S_j_ = spillover rate, H_i,j,t_ is hazard in grid cell *i* in year *t* as defined in steps 3 and 4 above, P_i,t_ = human population density in grid cell *i* in year *t* (from SSP populations^27^) and V_i,t_ = proportion of vulnerable people. Where H_i,j,t_ * P_i,t_ represents the expectation of P_i,t_ draws from binomial distribution with the probability of success H_i,j,t_ i.e. expected exposure = Binom(P_i,t_, H_i,j,t_).

We used the approximate spillover rate S_j_ to predict expected exposures and, therefore, weight higher-burden diseases relatively more in any summary, and it was calculated using present day annual cases taken from the literature *C_2015_* divided by the total human population in the present-day hazard area:

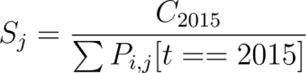

While the values for human populations P were taken directly from gridded data sources, we defined Vulnerability (V) as the proportion earning less than 25% of the median income which we calculated for present day and across five Shared Socioeconomic Pathways (SSP) scenarios^27^. We then used projected future Gini coefficients at the regional level to update present day per country Gini estimates^55^, to calculate the number of people projected to be earning less than a quarter of the median income, assuming that cumulative income distribution follows a parabolic curve and that poverty rate was geographically invariant. We then calculated mean values to summarise the differences in risk across continents, years and SSP scenarios. To account for variation across the dataset we expressed these results as the mean across 100 random bootstraps of 80% of the data, using the following notation:

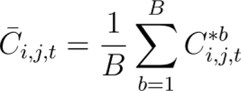

Where, *C_i,j,t_* represents the averaged estimate of the number of cases in grid cell *i* for agent *j* at time 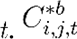 represents the bootstrapped estimate of cases for the *b*-th sample. *B* is the total number of bootstrap samples.

## Supplementary Data Folder

[link for preprint: https://www.dropbox.com/scl/fo/lrsts3ipcalppzktlol9e/h?rlkey=dnvh4rgpbe225pxdwwekpwq4z&dl=0]

## Data S1. (separate file)

Table of modelling parameters for all infectious agents examined and associated hosts and vectors.

## Data S2. (separate file)

Table of known locations of infectious agents found in two countries or less.

## Data S3. (separate file)

Locations of ten diseases that have point location data and sources of these data.

## Data S4. (separate file)

Table of regional and disease group summaries of burden trends by SSP scenarios

## Data S5. (separate file)

Table of climate models used in the future host/vector predictions

## Data S6. (separate file)

Zip file of estimated present day ranges with recorded locations.

---

**Figure S1.**
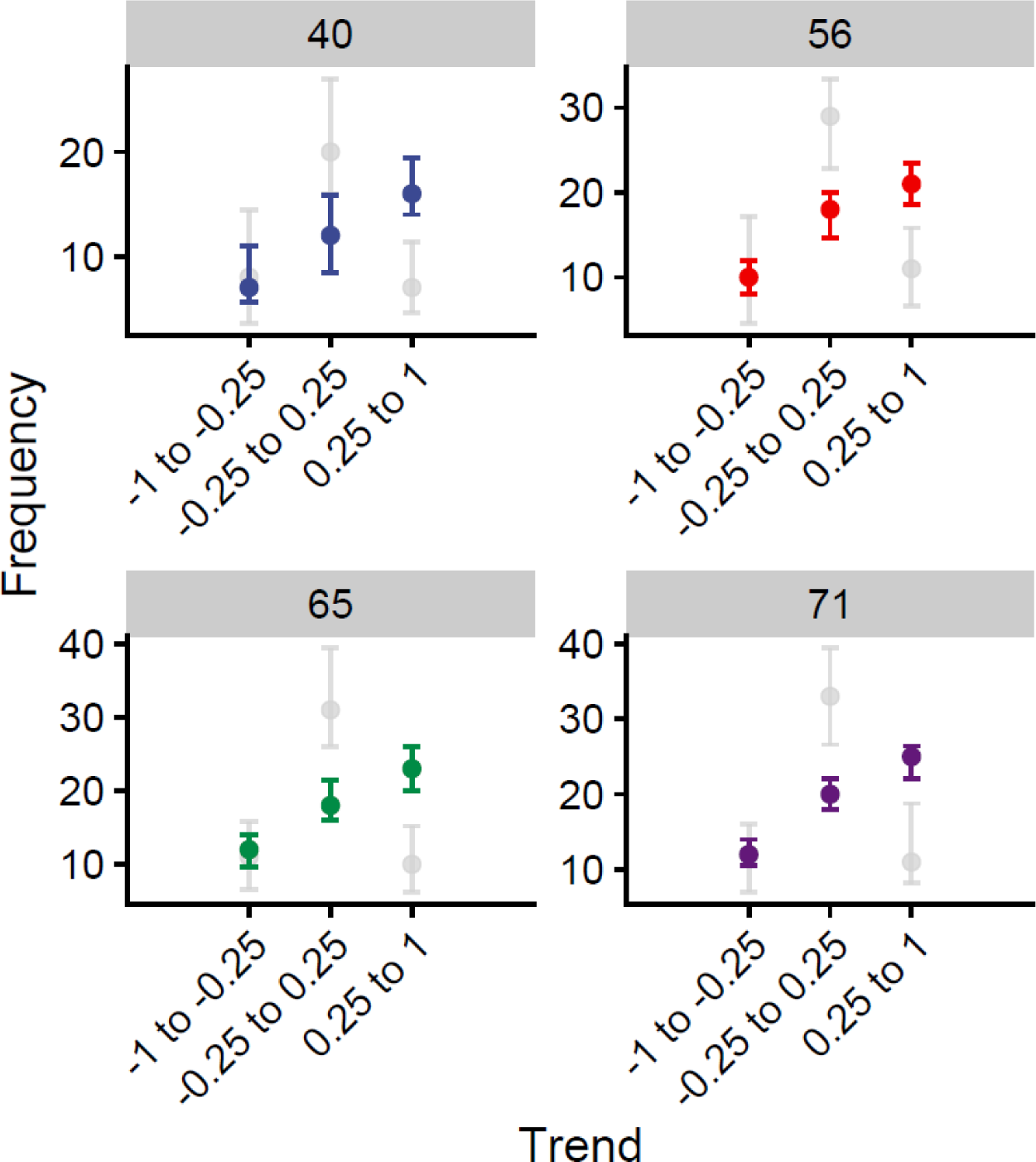
Variation in the response of zoonotic and vector-borne disease endemic hazards given climate change over the next 60 years (2030 to 2080). Each panel represents a different size subset of diseases (median *n* in panel banner) repeated taken at random 1000 times and the number of those disease (coloured dots) that had a spearman rank correlation between endemic area size and climate change (represented by year) of −1 to −0.25 (Contracting), −0.25 to 0.25 (Static) or 0.25 to 1 (Expanding). Whiskers represent 95% HPDI around the median. Grey dots and whiskers represent the same values but for 1000 simulations where endemic hazard size is randomised with respect to climate change.

**Figure S2.**
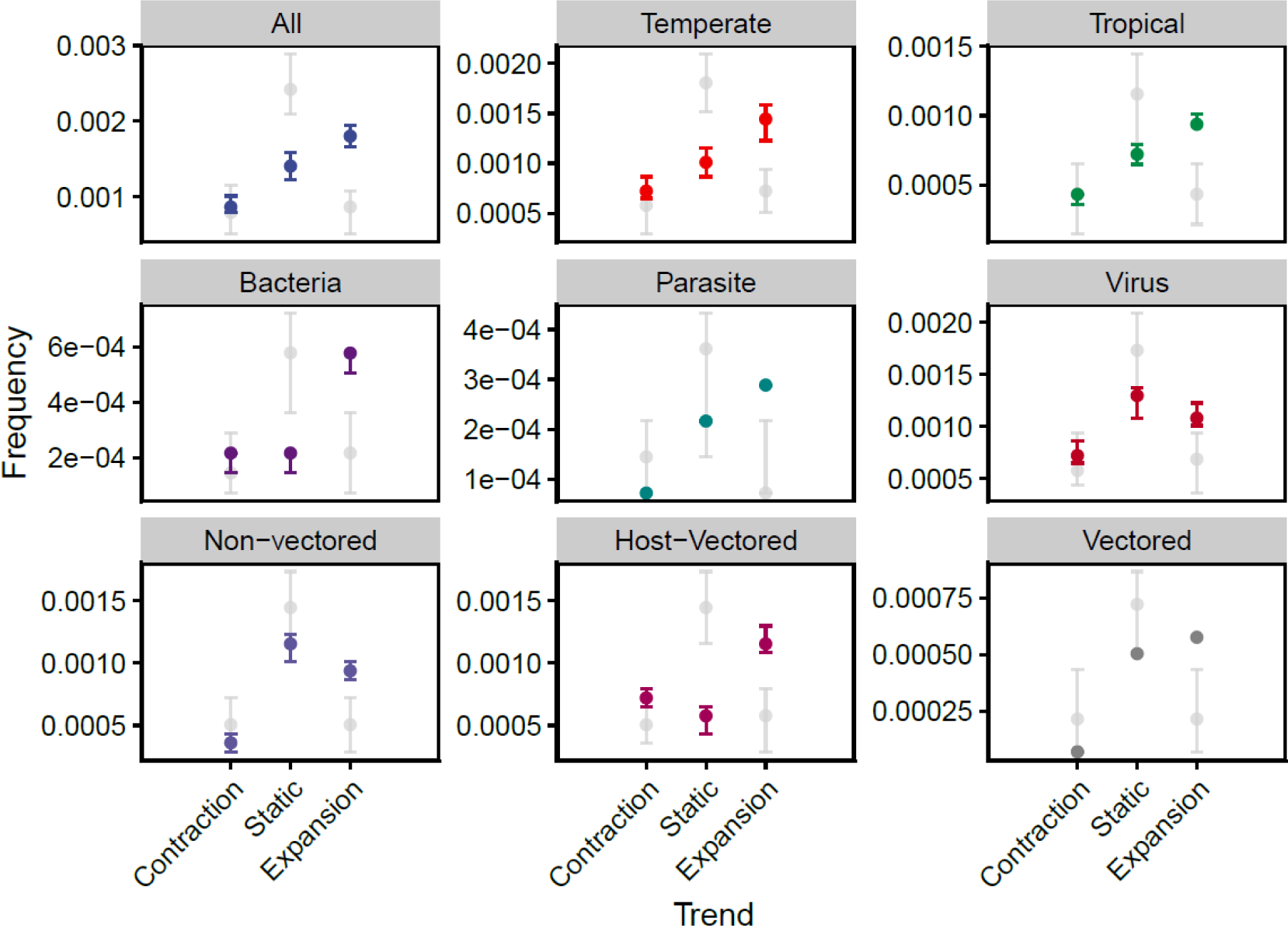
Proportion of disease endemic areas expanding, remaining static and contracting over time (coloured dots) compared to null expectation (grey dots). Variation in the response of endemic hazard size change of zoonotic and vector-borne diseases (n∼55 from 1000 random samples taken from an overall pool of 101 diseases) given climate change over the next 60 years (2030 to 2080). Coloured dots show the median relative frequency of diseases that had a spearman rank correlation of −1 to −0.25 (Contracting), −0.25 to 0.25 (Static) or 0.25 to 1 (Expanding). Whiskers represent 95% HPDI around the median. Grey dots and whiskers represent the same values but for 1000 simulations where for each disease year is randomised with respect to disease areas before the correlation is taken. Panels other than “All” represent the relationship in subsets of the data where only those diseases described in the panel title are included.

**Figure S3.**
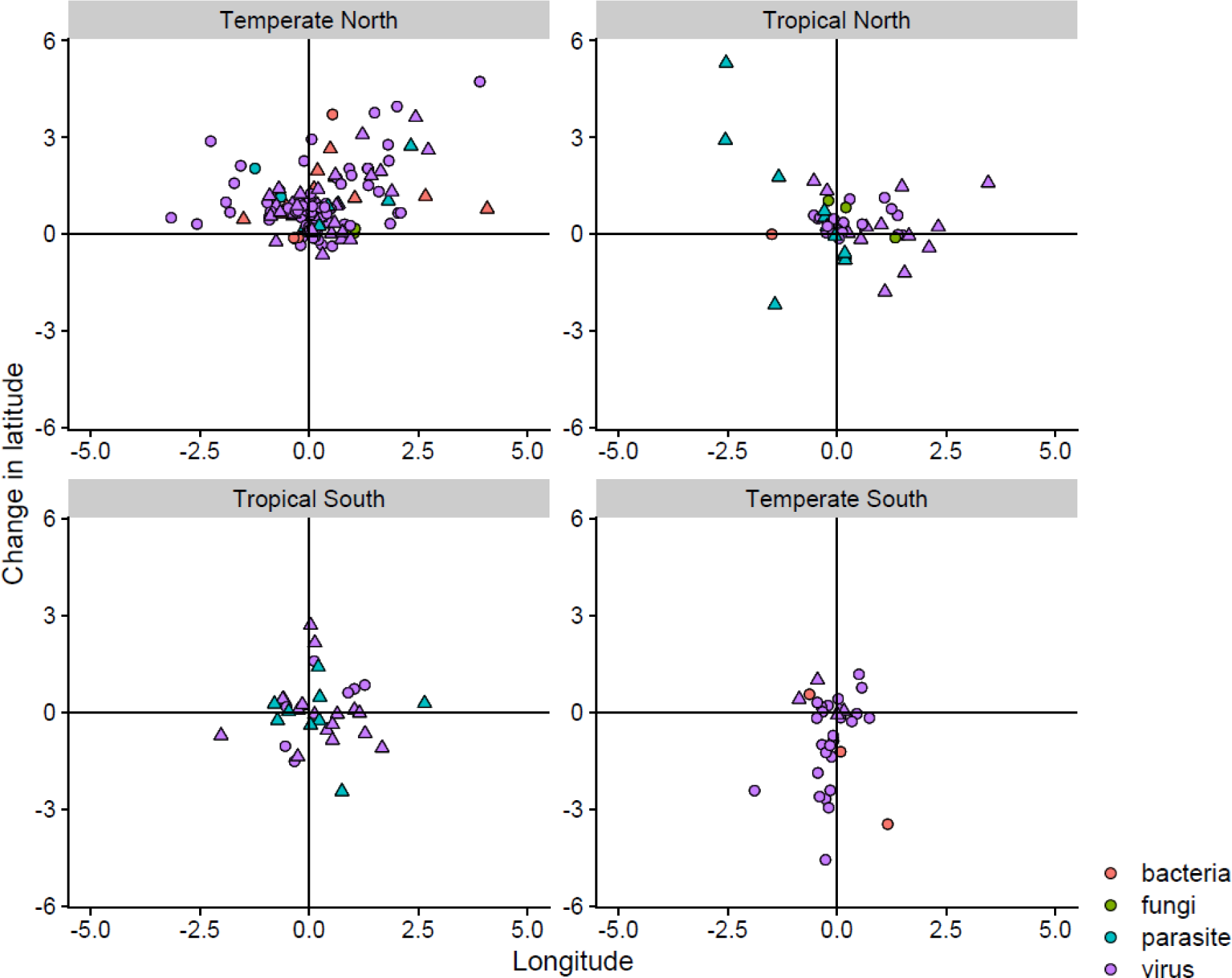
Expected change in degrees of the longitude and latitude of the centroid of 101 zoonotic and vector-borne disease endemic hazard limits from 2030 to 2080 The panel represent the global region the centroid belongs to, with temperate areas being greater than 23.5^ο^ north and less than −23.5^ο^ south and tropical areas from 0^ο^ to 23.5^ο^ and 0^ο^ to −23.5^ο^ for north and south tropical areas respectively. Triangles are vectored diseases whereas diseases represented as circles have no known vector. Colours represent the pathogen group.

**Figure S4.**
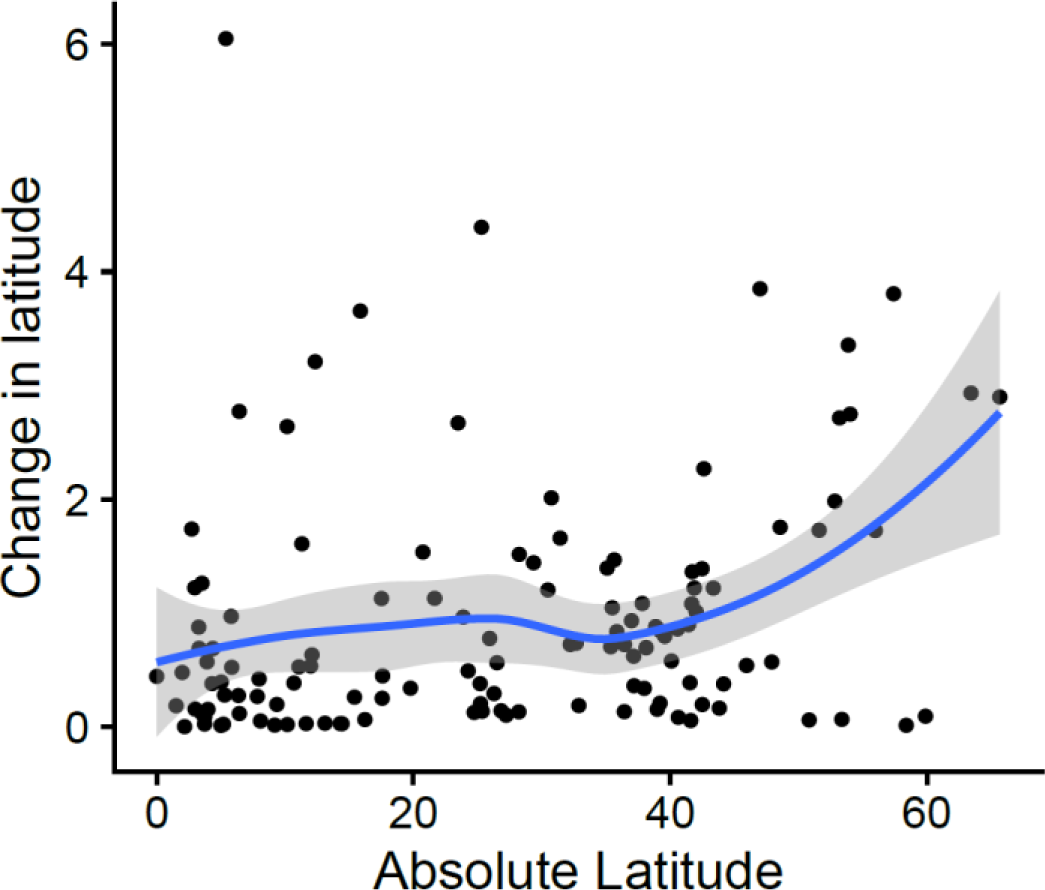
Relative latitudinal shift in degrees (y-axis) compared to the number of degrees from the equator (absolute latitude, x-axis). Dots represent change on each axis from 2015 to 2050 for individual disease projections and the line is the smoothed (loess) trend.

**Figure S5.**
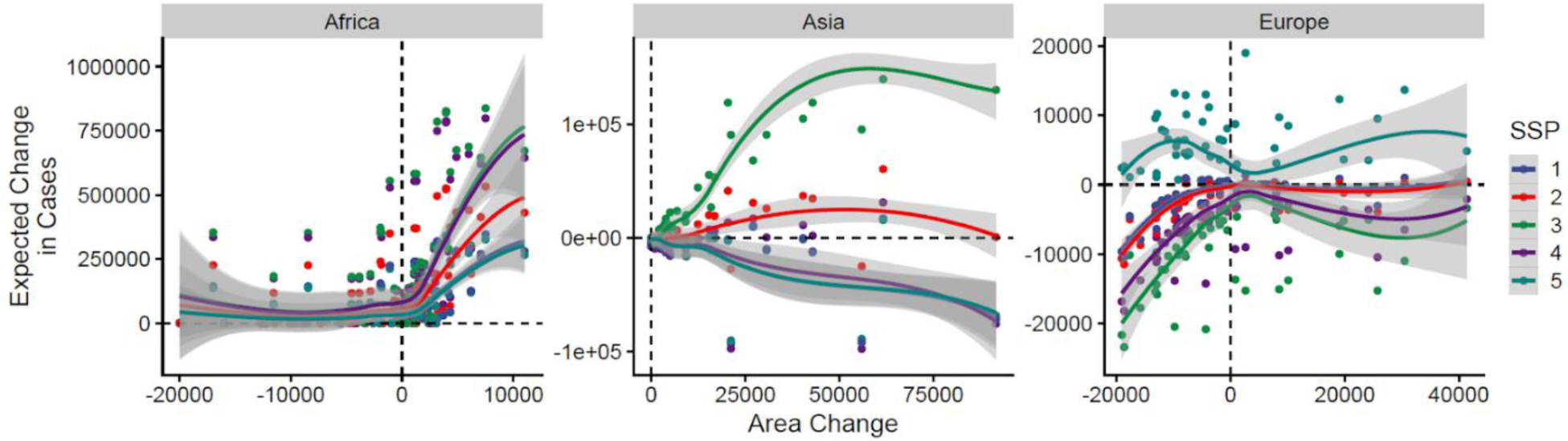
Relationship between change in disease endemic area from 2015-2050 and the expected change in disease cases over that same time period for RCP 4.5. Dots are individual per disease mean projections and lines are mean trends across all diseases. Colours represent different Shared Socioeconomic Development Pathways, defined in fig. 4.

**Figure S6.**
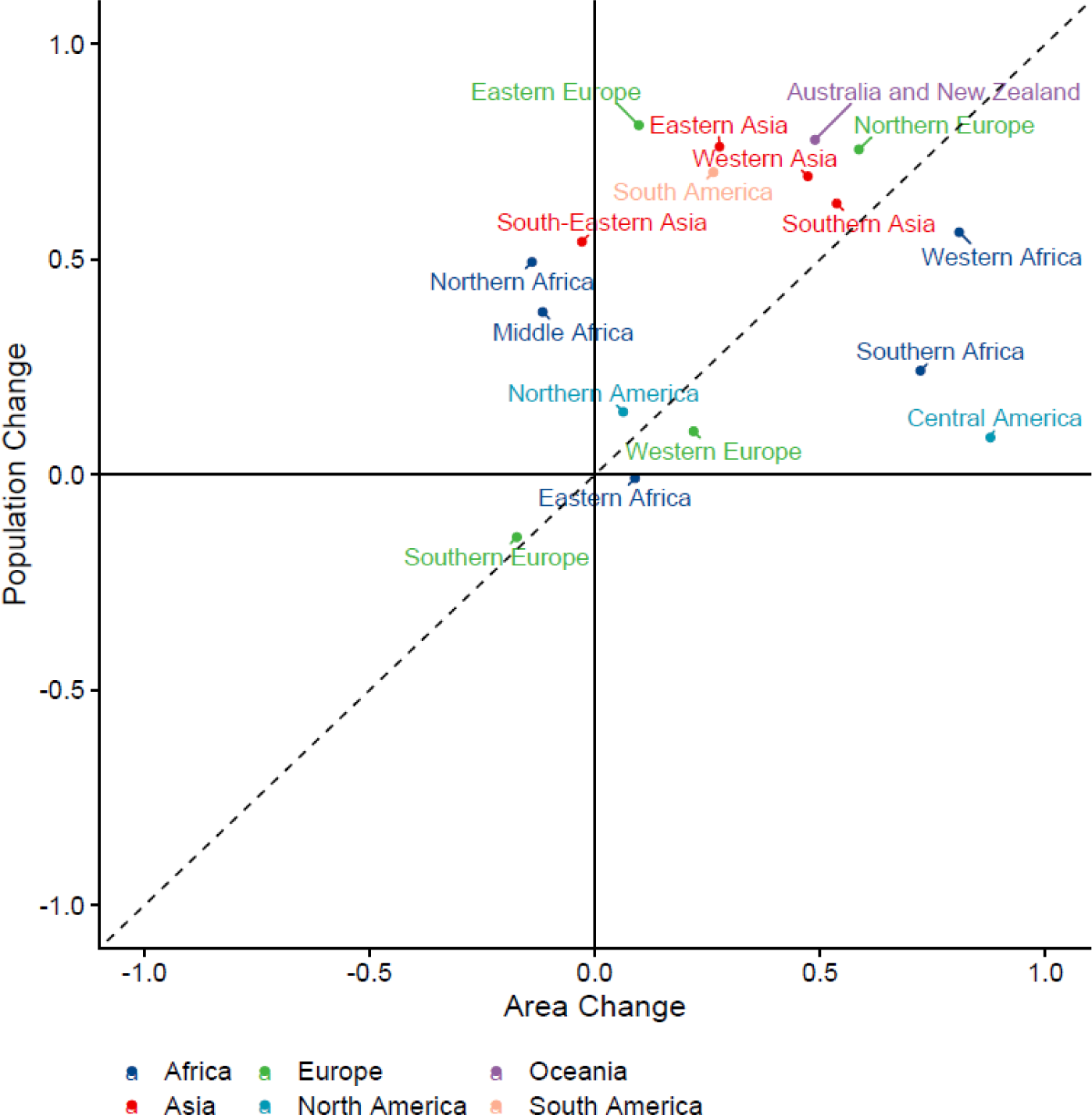
Influence of human population growth and climate change on size of area at risk of disease. Comparison of Pearson correlation of regional burden change and the change in human population density (y-axis), and Pearson correlation of regional burden change and the change in area at risk of disease due to climate change (x-axis), for zoonotic and vector-borne diseases whose endemic area centroid is located in different global subregions. All change is from the year 2015 to 2050 and for RCP4.5 and SSP2 (business as usual) and points are colour coded by continent.

**Figure S7.**
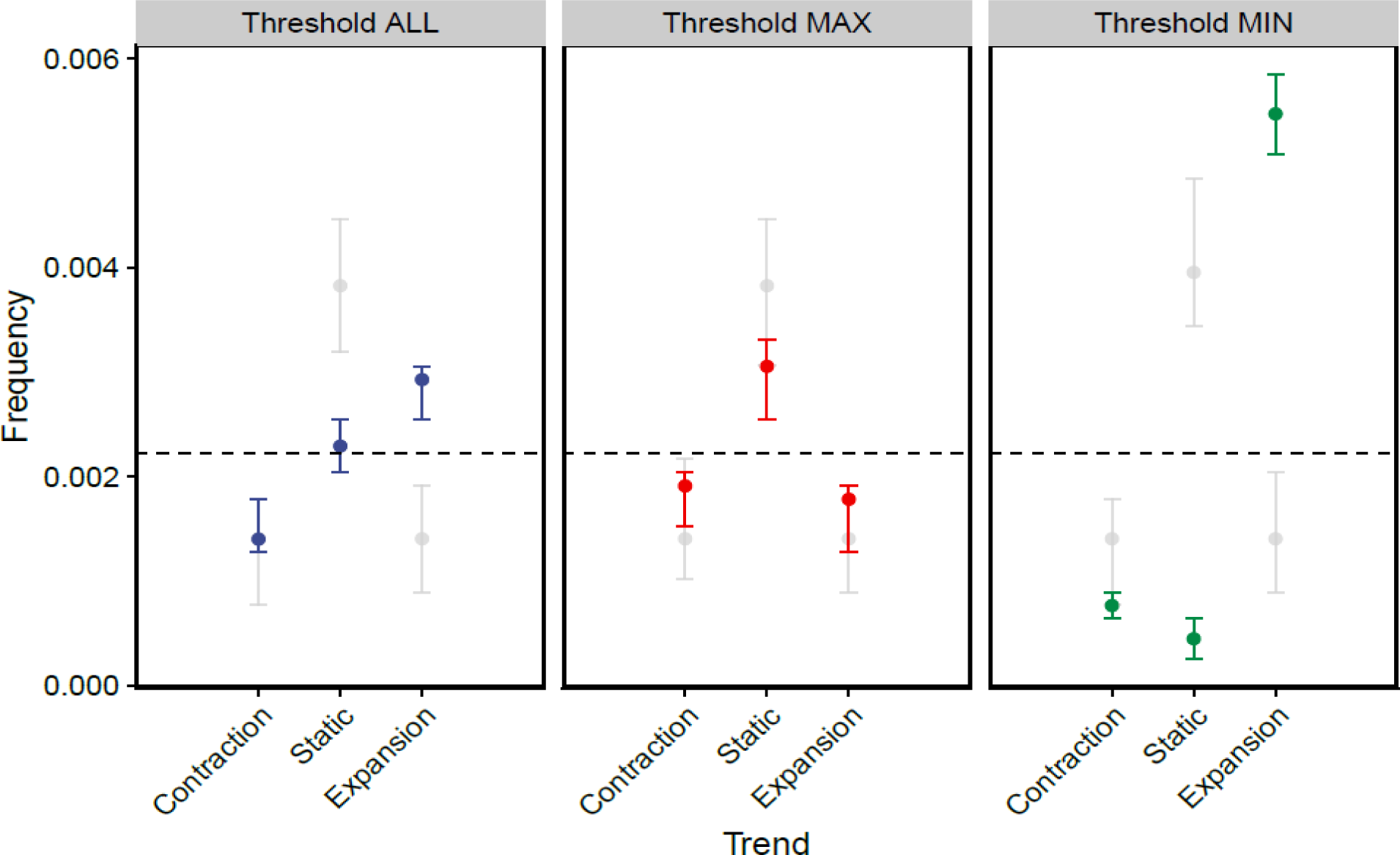
Variation in the response of zoonotic disease endemic hazards given climate change over the next 60 years (2030 to 2080). Each panel represents threshold used define the edges of the endemic hazards with panel (A) the mean best (max AUC) threshold, panel (B) the maximum possible threshold and panel (C) the minimum possible threshold and the number of those diseases (coloured dots) that were expanding, remaining static and expanding. Whiskers represent 95%HPDI around the median. Grey dots and whiskers represent the same values but for 1000 simulations where endemic hazard size is randomised with respect to climate change.

## Notes

### Competing Interest Statement

The authors have declared no competing interest.

